# Sunshine on KOL : a retrospective study about financial ties between medical key opinion leaders and pharmaceutical industry in France

**DOI:** 10.1101/2021.05.07.21255795

**Authors:** Marie Clinckemaillie, Alexandre Scanff, Florian Naudet, Adriaan Barbaroux

## Abstract

**Objective:** To investigate the nature, extent and evolution of financial relationships between key opinion leaders (KOL) or non-KOL physicians and pharmaceutical and device companies in France.

**Design:** Retrospective and descriptive study

**Setting:** All doctors practicing in France, with a focus on 548 KOL defined as board members of all the professional medical associations having published clinical practice guidelines in 2018 or 2019. These 99 associations were identified by the cross-checking of 3 databases.

**Main outcome measures:** The number and the amount of gifts (year by year since 2014), remunerations and agreements (year by year since 2017).

**Results:** Physicians had 818m€ ($936m, £741m) of gifts declared from 2014 to 2019. 83% of KOL had such links of interest. The 548 identified KOL represented 0.24% of physicians in France but received 1.5% of the total amount of gifts, i.e. €12.3m ($14m, £11m or €3 700 per capita per year).

Physicians had 125m€ ($143m and 114m£) of agreements declared from 2017 to 2019. The 548 KOL received 0.72% of the agreements and 2.5% of the value of the agreements, i.e. 3.1m€ ($3.6m, £2.8m) or €1 900 per capita per year ($2200, £1700).

Physicians had 156m€ ($178m and 141m£) of remunerations declared from 2017 to 2019. The 548 identified KOL received 4.4% of the total value of remunerations to physicians, i.e. 6.8m€ ($7.8m, £6.2m) or 4 100€ per capita per year ($4 800, £3 700).

Almost every professional medical associations (99%) had in their board at least one KOL with a financial tie.

**Conclusion:** Financial relationships between KOL and the industry in France are extensive, KOL have much more financial ties than non-KOL practitioners. The main limit of this study arises from the quality of information provided on the French Transparency in Healthcare database.

Pre-registration: osf.io/m8syh

**Strengths and limitations of this study:** This is the first attempt to provide data on the extent of the links of interest between opinion leaders and pharmaceutical industry in France.

Author crossed the nationwide databases of financial ties with three databases of professional medical associations.

All medical doctors practicing in France were inclused, with a focus on 548 KOL defined as board members of all the professional medical associations having published clinical practice guidelines in 2018 or 2019.

These 99 associations were identified by the cross-checking of 3 different catalogs of French professional associations.

The major links between key opinion leaders and industry ask the question of the independence of the experts, and raises concern that guidelines can be influenced by industry.

## INTRODUCTION

Financial ties between healthcare workers and pharmaceutical indusctry may affect every aspects of medical activity, from research to clinical practice. Clinical trials and meta-analyses sponsored by the pharmaceutical industry are more likely to conclude that drugs are effective than non-sponsored trials. ^1^ Industry’s transfers of value to physicians have been shown to be associated with more expensive, more frequent and of lower quality prescriptions^2–5^. Recommendations for clinical practice, which define the diagnostic criteria and treatment of the diseases, can also be under influence, since their authors often have ties with the industry. ^6–11^

Following the example of the USA with the US Physician Payments Sunshine Act, France created the Transparency in Healthcare public database (transparence.santé.gouv.fr) in 2014. ^12–14^ Pharmaceutical and medical device industries are required by law to disclose gifts, agreements and remunerations they transfer to healthcare professionals in France.

The term “Key Opinion Leaders” (KOL) refers to physicians who influence their peers’ medical practice, including but not limited to prescribing behaviour. It was coined by sociologists who demonstrated that people changed their opinions more because of some individuals in their networks than because of media or advertising: the influence of the physicians’ social networks is major to make them adopt a new drug. ^15,16^ Pharmaceutical companies hire KOLs at different stages of the drug development process, from clinical trials to promotion. ^17,18^ Typically, KOLs are physicians or researchers who are respected in their field and recognized for their work, such as broad members of professional medical associations. ^18–22^

Major ties between leaders of professional medical associations and the pharmaceutical industry have recently been described in North America. ^10,11^ In France, these links had never been studied yet.

In this study we described the nature and evolution of gifts, agreements and remunerations perceived by key opinion leaders (KOL) and other physicians using the data from the Transparency in Healthcare database. We also grouped gifts, agreements and remunerations perceived by these KOL for each professional medical association they belong to.

## METHODS

As per our protocol (registration number: osf.io/m8syh), we conducted a retrospective study of the financial relationships between industry and board members of the national professional medical associations publishing clinical practice guidelines.

### Identifying professional medical associations

Professional medical associations were defined as any group of physicians who published clinical practice guidelines in France. One author (MC) built the list of eligible associations by cross-checking three different databases: the “Catalogue et index des sites médicaux de langue française” (CISMEF) ^23^), “Le Parisien” review professional medical associations catalogue ^24^) and the “Bibliothèque Médicale AF Lemanissier” (BMLweb) ^25^). We included only national associations and excluded association titled as concerning “rare disease”. Then, MC searched for those who had published at least one clinical practice recommendation in 2018 or 2019 using Google scholar, academic medical library of the general hospital of Le Mans and CISMEF.

### Identifying Key Opinion Leaders

Using each professional medical association’s website, MC identified between October 2018 and May 2020 all physicians who were board members.

KOL were defined as members of the association’s board or governing council but not of sub-committees. KOL were identified by their name, medical specialty and city of practice, on the medical association website then if missing on google. Discrepancies and uncertainties were resolved by discussion with a second author (AB).

The Transparency in Healthcare database was downloaded on may 18, 2020 from the website EurosForDocs^26^. EurosForDocs is a tool inspired by the American website DollarsForDocs. EurosForDocs aims to help querying and understanding the Transparency in Healthcare database by cleaning and grouping payments by categories and beneficiaries. It also harmonizes the identification of doctors using their unique identification number in the National Healthcare Professional Registry : the “RPPS” (Répertoire Partagé des Professionnels de Santé). RPPS of the KOL were identified by AS from Health-Directory database and Transparency in Healthcare database. Uncertainties were resolved by manual inspection (MC).

### Identifying and extracting payment details

By using the RPPS unique identification number, data on payments for the identified leaders^27^ were extracted, using categories within the database: gifts, agreements and remunerations. We took into consideration the data from the date they were obligatory to declare: gifts from January 1, 2014 to December 31, 2019 and agreements and remunerations from January 1, 2017 to December 31, 2019.

“Gifts” include anything that is granted without consideration, in kind or in cash, directly or indirectly, of an amount greater than or equal to 10€ ($11,4) including taxes. “Remunerations” represent the payment by companies for work or services, of an amount greater than or equal to 10€. “Conventions” are agreements involving obligations on both sides: participation in a congress, research or clinical trial activity, training action, etc. The characteristics and date from where the payments were mandatory to declare are presented in **table 1**.

**Table 1.**
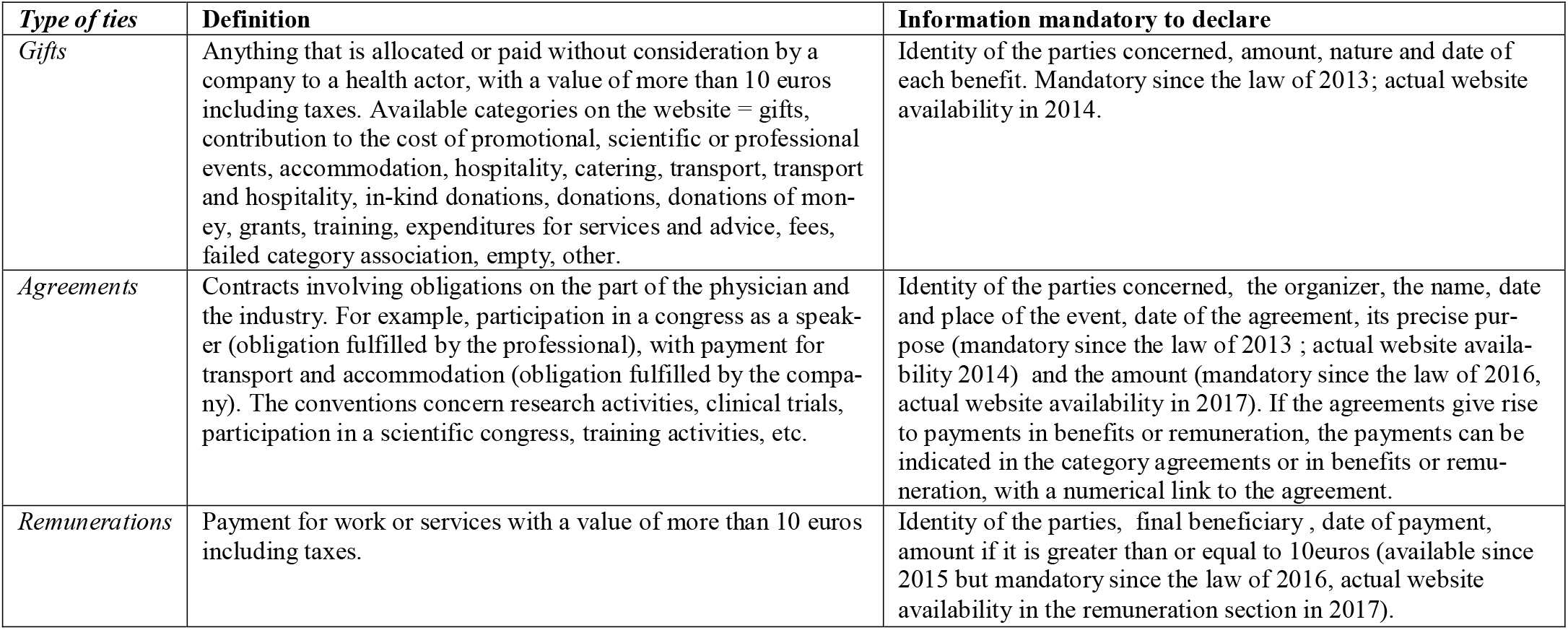
presents the 3 categories of links of interest and the date from where they had to be declared on the Transparency in Healthcare database (base Transparence Santé). The Transparency in Healthcare database was laid down in the “Strengthening the safety of medicines and health products” law of December, 2011, and launched in July, 2014.

### Outcome measures and descriptive analyses

The primary outcome was the total amount of gifts received by all the medical physicians and by the identified KOL, year by year since 2014.

A secondary outcome was the number and amount of the 2 additional categories of payments available after 2017 (i.e. agreements and remunerations), year by year since 2017.

Distribution of individual results of KOL pooled for each professional medical association is also presented. Quantitative data were described using median (inter-quartile range, IQR) rather than mean to be less biased by the influence of extreme observations. Binary outcomes were described using n (percentage). All analyses were performed using R. ^28^

### Changes to protocol

The secondary outcome including agreements and remunerations was not part of the protocol as these declarations were not mandatory before 2017. However, after having noted that remunerations represented more than 3 times the yearly amount of gifts, it was decided to include agreements and remunerations because we might have missed an important part of physicians-industry ties.

Then, as we identified some outliers with implausible amounts, it was likely that the database contained some errors (e.g. some gifts may have been reported in cents by the company [outliers typically ending in two zeros]). It was therefore decided a posteriori to exclude amounts exceeding 100 000€ ($118 000) for a single payment. It corresponds to 35 extreme observations (34 in 2019, 1 in 2018, i.e. 0.0005% of the gifts) and 32m€ (4% of the total and 13% of 2019).

### Patient and public involvement

Patients and public were involved throughout the French FORMINDEP association that aims to improve the independence of physicians’ medical education. FORMINDEP’s members (patients and physicians) kindly accepted to participate to the manuscript reviewing and editing. French CI3P organization (Patient and Public Partnership Innovation Center of the Faculty of Medicine of Nice) also accepted to participate to the manuscript reviewing and editing. Their comments enhanced the manuscript’s quality, especially the discussion.

## RESULTS

### Participants

We identified 238 professional medical associations. 101 of them had produced clinical practice guidelines in 2018 and/or 2019 and two of them had no website or no board on their website. We identified 605 KOL, 548 of them were found on the Transparency in Healthcare database. The number of KOL in each professional medical association ranged from 1 to 12, with a median of 6. 12 KOL belonged to more than one professional medical association. The way KOL were identified is described by the figure 1 : Flowchart.

**Figure 1.** Flowchart, representing how KOL were identified by crossing three databases.

### Transparency in Healthcare public database

The database contained 6b€ ($7.1b) of ties over 8 years. Gifts represented 1.7b€, agreements represented 1.3b€ and remunerations represented 3b€. ^26^ Gifts, agreements and remuneration are presented below from the year in which they were consistently declared, that is respectively since 2014, 2017 and 2017.

### Gifts (2014-2019)

For all physicians 7 354 492 gifts were declared for a total amount of 818m€ ($936m) from 2014 to 2019. The median amount for a gift was 46€ (IQR= 25-60, $54). Most KOL (83%) had at least one gift declared from 2014 to 2019. KOL’s gifts represented 0.68% of the number of all physicians’ gifts and 1.5% of the total amount of gifts, i.e. 12.3m€ ($14m). It represents a median of €3 700 of gifts per KOL per year. The median amount for a KOL’s gift was 60€ (IQR = 30–214).

Overall, the gifts declared to all physicians decreased in number and value from 1.3m gifts (151m€) to 923 000 gifts (108m€).

The number, value and proportion of gifts declared to KOL decreased from 9 687 gifts (0.70% of the total number of gifts to physicians) / 2.2m€ (1.5% of the total value of gifts to physicians) to 6044 gifts (0.65% of the total number of gifts to physicians) / 1.5m€ (1.4% of the total value of gifts to physicians).

The evolution year by year for each specific category of gift from 2014 to 2019 is presented in **Table 2**.

**Table 2.** Median (IQR) amount of gifts to KOL and non-KOL physicians in euros (€).

| *Gifts category* | *Physicians category* | 2014 | 2015 | 2016 | 2017 | 2018 | 2019 |
| --- | --- | --- | --- | --- | --- | --- | --- |
| *Accommodation* | *All physicians* | 210(165-314) | 215 (165-341) | 215 (170-314) | 215 (164-323) | 221 (170-349) | 218 (173-328) |
|  | *KOL* | 218 (172-305) | 220 (170-330) | 218 (179-296) | 226 (176-313) | 221 (175-323) | 229 (180-344) |
| *Hospitality* | *All physicians* | 50 (27-118) | 45 (25-60) | 46 (24-60) | 55 (30-60) | 53 (28-60) | 52 (28-60) |
|  | *KOL* | 70 (30-448) | 58 (28.2-258) | 60 (30-258) | 80.5 (40-362) | 60 (29-319) | 60 (30-329) |
| *Catering* | *All physicians* | 40 (24-55) | 40 (24-56) | 38 (23-55) | 38 (23-56) | 38 (23-56) | 38 (24-57) |
|  | *KOL* | 40 (23-59) | 40 (23-59) | 40 (23-58) | 40 (23-58) | 40 (24-59) | 40 (24-59) |
| *Transport* | *All physicians* | 208 (91-420) | 200 (88-398) | 191 (85-380) | 189 (79-357) | 182 (81-344) | 177 (77-332) |
|  | *KOL* | 202 (70-471) | 206 (74.2-461) | 244 (96-477) | 207 (77-445) | 198 (79.8-414) | 187 (71-395) |
| *Contributions to the cost of promotional events* | *All physicians* | 60 (50-400) | 60 (50-404) | 60 (44-400) | 200 (55-455) | 400 (250-590) | 440 (260-650) |
|  | *KOL* | 320 (60-591) | 350 (60-600) | 390 (87-650) | 450 (186-729) | 470 (290-740) | 538 (290-796) |
| *Donations-Grants-Training* | *All physicians* | 21 (17-30) | 23 (17-50) | 36 (23-94) | 62 (30-171) | 55 (29-171) | 55 (25-144) |
|  | *KOL* | 83 (60-275) | 68 (27.5-1578) | 96 (63.5-138) | 76 (45-192) | 155 (42-225) | 80 (42-180) |
| *Service and consulting* | *All physicians* | 26 (22-30) | 30 (22-45) | 30 (24-50) | 30 (25-40) | 50 (25-80) | 116 (40-362) |
|  | *KOL* | 30 (24-120) | 47 (29-325) | 65 (30-600) | 32 (25-83) | 130 (74-309) | 158 (69-506) |
| *Other* | *All physicians* | 100 (30-350) | 140 (30-496) | 100 (19-375) | 22 (16-84) | 25 (16-104) | 49 (16-220) |
|  | *KOL* | 337 (34.8-1000) | 800 (195-1188) | 700 (100-1000) | 310 (23-915) | 40 (12-153) | 79.5 (12-500) |
| *TOTAL* | *All physicians* | 45 (25-60) | 45 (25-60) | 45 (25-60) | 46 (25-60) | 48 (25-60) | 49 (26-60) |
|  | *KOL* | 59 (29-198) | 60 (30-224) | 60 (30-217) | 60 (30-213) | 60 (31-214) | 60 (31-210) |

Almost all (99%) associations had at least one member of its board who had at least one declared gift since 2014. The median amount of gifts declared for all the corresponding KOL of a professional medical association was 61 000€ (IQR= 14 000-143 000 ; $70 000) but varied widely between associations. 1% of the associations had no gift declared for their KOL, 16% had less than 1 000€ gifts per year for their KOL. 39% had between 10 000€ and 50 000€ gifts and 11% had more than 50 000 € gifts declared for their KOL each year.

### Agreements (2017-2019)

Concerning non-KOL physicians, 1.67 millions agreements were declared for a total of 125m€ ($143m) from 2017 to 2019. There were 1.28 millions agreements (77%) for which the reported amount was null. A null amount can be explained either by a report in one of the two other categories (when the agreement is linked with a gift or remuneration), or by a wrong declaration.

KOL’s agreements represented 0.72% of all agreements declared to physicians and 2.5% of the value of these agreements, i.e. 3m€ ($3,6m). It represents a median of €1 900 of declared agreements per KOL per year. There were 9 496 KOLs’ agreements (79%) for which the reported amount was null.

Overall, agreements declared to all physicians were increasing from 42m€ in 2017 to 43m€ in 2019.

The evolution year by year of the total amount, and median amount of agreements is presented in **table 3**.

**Table 3.**
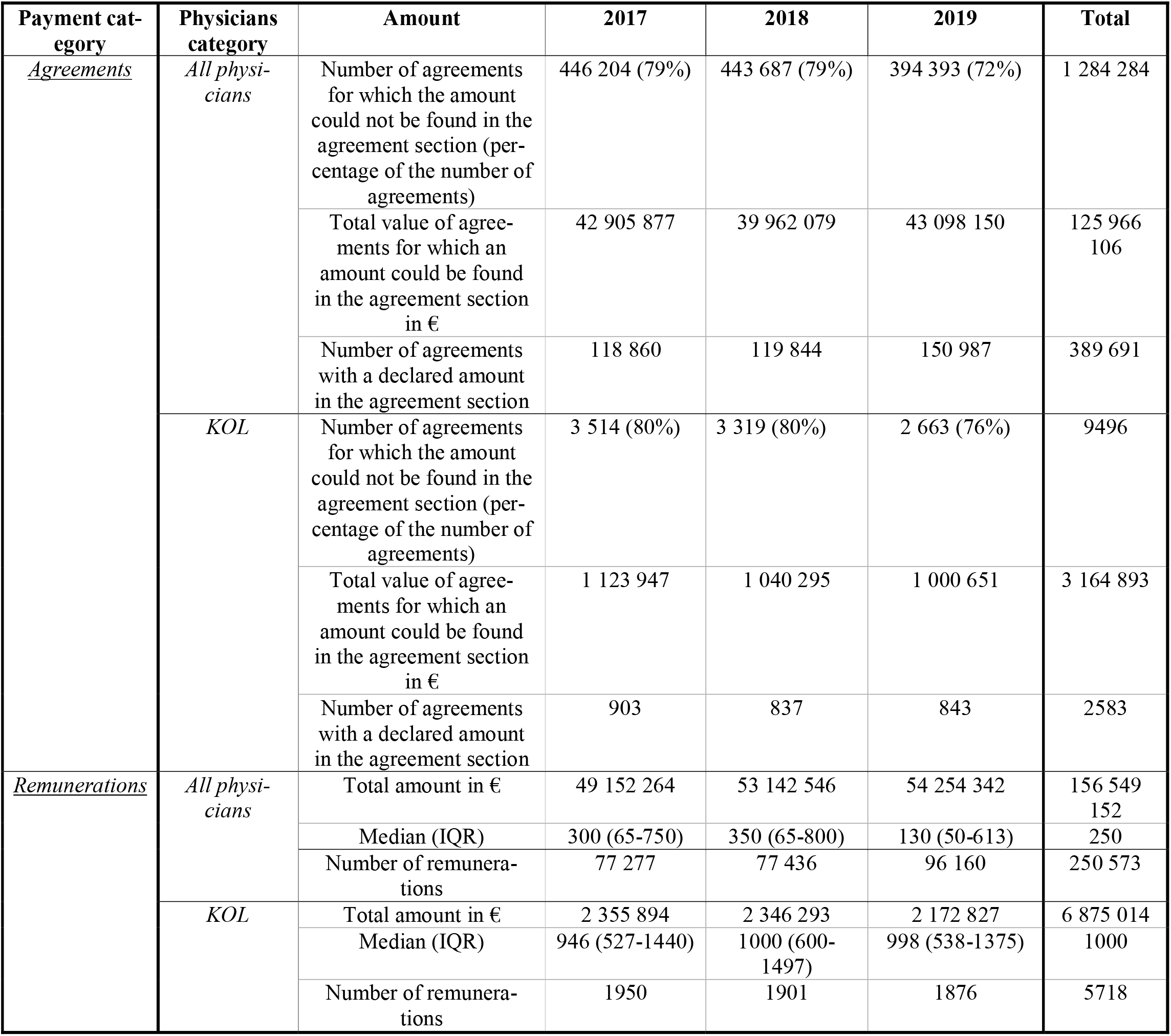
Total amount, and median (IQR) amount of agreements, total and median (IQR) amount of remunerations to KOL and non-KOL physicians, year by year since they are consistently declared.

The number, the value and the proportion of agreements declared to KOL decreased each year, from 4 400 agreements (0.78% of the number of agreements to physicians) / 1.1m€ (2.6% of the value of agreements to physicians) in 2017 to 3 500 agreements (0.64% of the number of agreements to physicians) / 1m€ (2.3% of the value of agreements to physicians) in 2019. This evolution is depicted in **Figure 2**.

**Figure 2.** Evolution of the 3 kinds of financial ties for KOL and all physicians. The gifts declared to all physicians were decreasing in number and value over time; the number, the value and the proportion of gifts declared to KOL were decreasing. The agreements declared to all physicians were increasing; the number, the value and the proportion of agreements declared to KOL were decreasing. The remunerations declared to all physicians were increasing; the number, the value and the proportion of remunerations declared to KOL were decreasing.

The median amount of agreements declared to all the corresponding KOL of an association was 15 900€ per year (IQR= 390 to 35 617).

#### Remunerations (2017-2019)

For all physicians, 250 873 remunerations were declared totaling 156m€ ($178m) from 2017 to 2019. The median amount for a remuneration was 250€ (IQR 55-742) ($296). KOLs’ perceived 2.3% of physicians’ remunerations, i.e. 6.8m€ ($7.8m) or 4.4% of the total value of remunerations to physicians. Overall, KOLs received 4 times more remunerations than other physicians, which represents a median of €4 100 of remunerations per KOL per year.

Regarding all physicians, remunerations increased in number and total value but the median amount decreased sharply. The evolution of the total amount of remunerations is presented in **table 3**. Physicians’ remunerations increased from 77 277 remunerations / 49m€ in 2017 to 96 160 remunerations / 54m€ in 2019.

The number, value and proportion of remunerations declared to KOL decreased each year from 2017 (1 900 remunerations, 2.5% of the number of remunerations to physicians, accounting for 2.3m€ and 4.8% of the value of remunerations to physicians) to 2019 (1 800 remunerations, 1.9% of the number of remunerations to physicians, accounting for 2.1m€ and 4% of the value of remunerations to physicians in 2019).

The median amount of remunerations declared for all the corresponding KOL of an association was 21 000€ per year (IQR 1012 - 68 977, $25 000).

## DISCUSSION

### Principle findings

During this period, 818m€ of gifts, 125m€ of agreements and 156m€ of remunerations were declared to physicians in France. The amount of gifts decreased and the total amount of declared agreements and remunerations increased. Gifts represented the largest amount declared.

Almost every professional medical association included at least one KOL who received one or more gifts since 2014 (99%) or 2017 (97%). Over the whole period, the median amount of gifts per association was €61 000 ($70 000). From 2017 to 2019, the median cumulative amount for each professional medical association was 15 900€ of agreements and 21 900€ of remunerations. The number and amount of gifts varied widely from one association to another, ranging from €0 to 160 000€ ($189 000) for all the members of one association on the studied period.

The number, value and proportion of gifts, agreements and remunerations for KOLs were slightly decreasing over time. Remunerations represented the largest amount declared to KOLs with a median amount per capita 4 times higher than for other physicians. KOLs represented 0.24% of the physicians but were associated to 1.5% of the gifts, 2.4% of the agreements and 4.4% of the remunerations in value. It represents €3 700 of gifts, €1 900 of agreements and €4 100 of remunerations per capita per year. This amount of agreements is probably underestimated since 79% of KOLs’ agreements amount was declared null in the database (see above).

### Strengths and Limitations

This study is exhaustive of all ties declared on the French Transparency in Healthcare database. All physicians practicing in France were included since ties are mandatory to declare.

However, the statements may be underestimated since many agreement’s amounts were not available. Indeed, when a physician signs an agreement conferring an advantage, the amount can be declared either nil, in agreement, in gift or both in agreement and gift. There is no government control at this level.

The effect of this bias is difficult to predict : on the one hand, firms did not declare the amounts of thousands of agreements, thus underestimating the amounts received by physicians. On the other hand, the amount of an agreement could be double counted. The entire Eurofordocs database (with all beneficiaries, without time limitations) contains 5.5 million agreements, 3.3 million of which have a nil amount. 2.2 million gifts claim to be linked to an agreement, but have an invalid textual link. There are therefore at least 1.1 million agreements with a nil amount despite the legal obligation to declare it.

Another limitation lies in the fact that the data comes from the declarations of the pharmaceutical industry itself with typos. Moreover, there may be a delay in data reporting, and remunerations may have been misclassified as it was possible to declare them as gifts or as remunerations until October, 2017.

Finally, in the absence of an official definition, we choose an objective but restrictive definition of KOL which lead us to rule out many individuals of great leverage that could also be called KOL.

### Comparison with other studies

Our results are in line with those observed worldwide but adds new data regarding the French context. Very recent U.S. study showed that nearly three-quarters of the executives of the 10 most influential professional medical associations in the U.S. had ties with the pharmaceutical industry, with wide variations in the amount of payments reported between the professional medical associations. ^10^ Total general payments of $24.8m (20.8m€, £18.9m) were linked to the 235 KOL of the 10 most influential professional medical associations in 3 years. The total median general payment was $6 000 (IQR $309 to $54 000) (5000€, £4 500).

In this study, KOLs received 10 times more per capita per year in total amount than the French KOLs, and more than 83 times more in terms of median amount.

The amount of money involved in this American study seems to be much more important. This difference could be explained by societal differences but also by the fact that we included professional medical associations regardless of their size, cost or influence. On the other hand, this difference can be explained by the fact that USA represents a population 5 times larger and 4 times more physicians, which may constitutes an important return on investments. Finally, in the US, there are more mandatory payments to report, and there are enforcement measures and effective penalties.^29^

### Implications of this study

Despite multiple calls for more distance,^10,30–33^ KOL have still privileged relationships with pharmaceutical industry. This phenomenon can lead to lower guidelines’ quality and to a general loss of confidence in both KOL and physicians. Indeed, several guidelines were abrogated since there have been doubts about the independence of the experts involved in their writing. ^34–37^ In turn, Chakroun et al. have shown that conflict of interests disclosure reduces public and physicians’ trust in KOL.^38^ Experience shows that financial ties can also be instrumentalized to discredit any expert position, the link being used as an argument to call into question the scientific opinion. ^39–41^

Our study’s finding of remaining concealment of the agreements amounts, despite the legal obligation to declare them, shows that transparency is still in progress and that both researchers and citizens do not yet have access to all data. For us, the main area for improvement would be to make it mandatory to report the amount of benefits and remunerations conferred by the agreement in the agreement section. Moreover, the declarations should be checked by the public authorities, which is the only guarantee of the reliability of the information provided.

Future research might focus on the correlation between the amount of gifts and the medical specialty or the cost of the concerned diseases. Further research is needed to identify other kinds of KOL such as the department heads of the teaching hospitals, and the medical university lecturers. Financial ties could be tracked over time, acting as a nudge to help chart moves towards independence.

## Data Availability

Data from EurosForDocs are available on https://www.eurosfordocs.fr/data#donn-es. Analytic code from the study is available on https://osf.io/4756p/?view_only=df16d649d87847e5aa9478960620bf81

## Acknowledgments

We thank Pierre-Alain Jachiet for his proofreading of the manuscript, his help to fully understand the database and use properly EurosForDocs.

## Footnotes

### Author Contributions

MC and AB initiated and designed the study, searched the literature, interpreted the results and wrote the manuscript. AS performed the analysis, contributed to the study design and interpreted results. FN contributed to the study design and interpreted the results. AB is the garantor. All authors have critically revised the manuscript and approved the manuscript. The corresponding author attests that all listed authors meet authorship criteria and that no others meeting the criteria have been omitted.

### Ethical approval

the French Commission Nationale Informatique et Libertés approved this study.

### Data sharing statement

The guarantor (AB) affirms that the manuscript is an honest, accurate, and transparent account of the study being reported; that no important aspect of the study was omitted; and that any discrepancies from the study as originally planned have been explained.

